# Semi-Supervised Learning in Prostate MRI Tumor Segmentation Approaches Fully-Supervised Performance on External Validation

**DOI:** 10.1101/2025.05.13.25327456

**Authors:** Eduardo H.P. Pooch, Georgios Agrotis, Lishan Cai, Mark Emberton, Taimur T. Shah, Hashim U. Ahmed, Regina G.H. Beets-Tan, Sean Benson, Tomas Janssen, Ivo G. Schoots

## Abstract

**Purpose:** To evaluate the diagnostic performance of semi-supervised learning models for aggressive prostate cancer segmentation on MRI compared to fully-supervised models trained with additional expert annotations.

**Materials and Methods:** We used 1500 MRI scans from the PI-CAI challenge training subset. Positive scans had 220 human and 205 AI-generated annotations. The mtU-Net (proposed teacher-student semi-supervised approach) was compared to supervised (trained using only 220 human annotations) and semi-supervised (trained on human and AI-generated annotations) nnU-Net. The 205 AI-annotated scans were manually annotated, and a fully-supervised model was trained. External validation was performed on a newly annotated dataset from the PROMIS study (n=574) and the Prostate158 dataset (n=158). Patient-level performance was evaluated using Area Under the Curve (AUC), Average Precision (AP) for lesion-level detection, and the DeLong test to compare performance.

**Results:** The fully-supervised nnU-Net showed the highest performance on the internal PI-CAI test set (AUC=0.89[0.87-0.91]/AP=0.65[0.60-0.70]) and external validation datasets PROMIS (AUC=0.70[0.66-0.74]/AP=0.24[0.19-0.29]) and Prostate158 (AUC=0.87[0.82-0.92]/AP=0.64[0.56-0.72]), significantly outperforming the supervised baseline (p≤0.002). The proposed semi-supervised mtU-Net demonstrated close external validation performance on PROMIS (AUC=0.66[0.62-0.71]/AP=0.20[0.16-0.25]) and Prostate158 (AUC=0.86[0.81-0.92]/AP=0.58[0.49-0.67]), significantly outperforming the supervised baseline on both datasets (p=0.024 and p=0.007, respectively). Semi-supervised nnU-Net showed intermediate results on PROMIS (AUC=0.65[0.60-0.69]/AP=0.20[0.16-0.24]) and Prostate158 (AUC=0.81[0.74-0.88]/AP=0.53[0.44-0.62]), significantly outperforming the supervised baseline only on PROMIS (p=0.042).

**Conclusion:** In prostate MRI tumor segmentation, nnU-Net fully-supervised learning performed best. However, in external validation, mtU-Net’s semi-supervised learning performance approached the fully-supervised model, demonstrating a valuable approach when expert annotations are limited.

**Summary:** Semi-supervised learning achieves close performance to fully-supervised methods on external validation in prostate cancer segmentation, reducing dependence on expert annotations in increasing demands.

**Key points:**

- The inclusion of AI-annotated data during training showed close performance to annotating additional samples with expert delineations, suggesting data diversity may be as impactful as increased expert annotation volume.
- The combination of pseudo-labeling with consistency regularization within the semi-supervised mtU-Net framework mitigated the impact of potential inaccuracies in AI-generated annotations, resulting in performance approaching that of fully-supervised models.

## 1. Introduction

Prostate MRI has become a pivotal tool in early cancer detection, accurate diagnosis, and effective management [1–3]. However, the reader-dependent nature of identifying aggressive cancer (Gleason Grade Group ≥ 2) on MRI leads to significant variability across centers and an overall low positive predictive value [4]. Deep learning methods may improve manual analysis and interpretation, enhancing MRI accuracy [5].

A significant obstacle in training deep learning models for medical image segmentation is the need for manually voxel-level annotated datasets by radiological experts. This process is time-consuming and costly. In the era of increasing demand for annotated data, semi-supervised learning (SSL) offers a solution by leveraging manually labeled and unlabeled data, reducing dependence on fully-annotated datasets. A common SSL technique is pseudo-labeling [7], using predictions from a pre-trained supervised model to generate labels for unlabeled data, effectively expanding the dataset for further training.

Previous SSL studies in medical image segmentation have primarily focused on relatively simple tasks, such as anatomical structure segmentation and tumor segmentation, where tumor presence is consistent across all samples [8, 9, 10]. These tasks can often achieve high performance with a few fully labeled samples (e.g., 20–50) with established segmentation methods like nnU-Net [11]. These SSL studies often simulate limited-label scenarios by using only a small subset of the available annotations in the dataset.

In contrast, segmenting aggressive tumors in a diagnostic cohort is considerably more complex than the tasks typical of previous SSL studies. This increased difficulty arises because tumors often occupy a small portion of the image, have irregular shapes, and may not be present in many patients within this cohort, requiring a different approach than standard segmentation tasks. The PI-CAI challenge [6] is a machine learning competition to validate algorithms for detecting aggressive cancers on prostate bi-parametric (bp) MRI. It provided a platform for developing and comparing machine learning solutions, focusing on semi-supervised approaches.

This study systematically compares the performance of supervised and semi-supervised learning approaches for prostate cancer segmentation in a large-scale training and validation setup. We hypothesize that SSL can achieve comparable performance to supervised models trained with fully-annotated datasets. Furthermore, this study investigates the practical applicability of SSL to a real-world clinical challenge and its generalization to external datasets.

## 2. Methods

### 2.1. Data

We used the data made available for the PI-CAI challenge [6], consisting of 1500 bi-parametric MRI (bpMRI) scans for training and two hidden test sets, Test 1 with 100 samples from the same centers as training and Test 2 with 1000 hidden test samples, including 197 samples from one external center. The training set included 425 GG≥2 positive, of which 220 were human-annotated, while the remaining 205 samples were AI-annotated. The remaining 1045 cases were negative for GG≥2 lesions. To evaluate the effect of increasing the number of human annotations, a prostate radiologist with 8 years of experience manually annotated the 205 AI-annotated images from the PI-CAI challenge dataset. These annotations were based on the bpMRI sequences available, and the radiologist was aware of the number of GG≥2 lesions reported in histopathological findings. The annotations were made on ADC using the 3D Slicer software (version 5.0.3), and the original bpMRI sequences remained unaltered. The expert annotations performed for this study will be made publicly available.

One limitation of the PI-CAI [6] test sets is that most samples are obtained from the same centers as the PI-CAI training data. Therefore, we conduct external validation using two independent datasets, PROMIS [13] and Prostate158 [14], with a total of 732 scans to assess the generalizability of SSL-based segmentation models. PROMIS is a multicenter confirmatory study from the UK. All 574 prostate cancer-suspected men in the study underwent 1.5 Tesla MRI and transperineal biopsy with a sampling interval of 5mm. This saturation biopsy approach ensures a thorough examination of the prostate tissue, addressing the limitation of potential undersampling often associated with less rigorous biopsy techniques, such as targeted or systematic. Based on the available biopsy reports, we performed voxel-level annotations to manually delineate 565 cancerous lesions on their ADC maps; out of those lesions, 396 were GG≥2. The tumor annotation on bpMRI was based on the histopathological report, ensuring the annotated lesions have a strong ground-truth for GG≥2 presence.

Prostate158 [14] is an open-source dataset specifically curated for developing and evaluating algorithms in prostate cancer research. It comprises 158 patient cases with 3T bpMRI scans of the prostate gland, with expert annotations of anatomical zones and cancerous lesions. The dataset includes T2-weighted and diffusion-weighted sequences with apparent diffusion coefficient maps. Histopathologic confirmation is available for all identified cancerous lesions.

To visualize the typical locations of lesions identified by each annotation source (AI-generated, human-annotated, and pathology-confirmed from PROMIS), binary lesion masks were aggregated across all patients within each respective group. These aggregate 3D distributions were then projected onto a 2D plane by summing voxel presences along the z-axis, creating the heatmaps shown in Figure 3. Relative coverage was computed based on the 3D volume covered by the AI annotations and the volume covered by the human annotations. These volumes were then expressed as percentages relative to the total 3D volume covered by the union of both masks. The 3D Intersection over Union (IoU) was calculated between the complete AI-generated segmentation mask and the complete human-annotated segmentation mask for each of the 205 patients. The mean of these 205 patient-level IoU values was reported to summarize overall spatial agreement per case. To assess the degree to which AI annotations localized individual human-annotated lesions (used as the reference standard for this specific comparison), a lesion-level hit ratio was calculated. A ‘hit’ was defined as any individual human-annotated lesion achieving a 3D IoU of 0.10 or greater with any overlapping region from the corresponding AI-generated mask. The hit ratio was calculated as the total number of ‘hit’ human lesions divided by the total number of human-annotated lesions.

### 2.2. Evaluation Metrics

Model performance was evaluated using the PI-CAI challenge score [6], the average of two metrics targeting distinct tasks: Area Under the Receiver Operating Characteristic Curve (AUC) for patient-level diagnosis and Average Precision (AP) for lesion-level detection. For AUC computation each patient scan was assigned the highest voxel prediction score indicating the likelihood of containing aggressive (GG≥2) cancer. The AUC was derived from the Receiver Operating Characteristic (ROC) curve analysis, summarizing the model’s ability to discriminate positive from negative cases overall without selecting a specific threshold. ROC curves for external validation are shown in Figure 4. The AP computation focused on lesion-level detection and it represents the area under the precision-recall curve. A predicted lesion detection was classified as a True Positive (TP) based on an IoU>0.10 with a corresponding ground-truth lesion annotation. Predictions not meeting the hit criterion were classified as False Positives (FP). AP was then calculated based on the precision and recall values derived from ranking all predicted lesion detections, selecting the highest lesion voxel score as the confidence and applying the TP/FP classification. 95% confidence intervals (95% Cis) for both AUC and AP were determined using bootstrapping with 10,000 iterations. The statistical significance of differences in AUC between each strategy and its corresponding supervised baseline was determined using the DeLong test, with p<0.05 considered significant.

### 2.3. Supervision strategies

A supervised learning model was trained using the 220 human-annotated samples. This supervised model was used to annotate samples identified as GG≥2 positive based on the information on the reports, generating 205 AI-labelled samples [12], which were included in the PI-CAI challenge training set. These AI-labelled samples were used for the semi-supervised approaches. A second supervised model (referred as fully-supervised) was trained on newly annotated data by a radiologist on these 205 samples (Figure 1).

**Figure 1:**
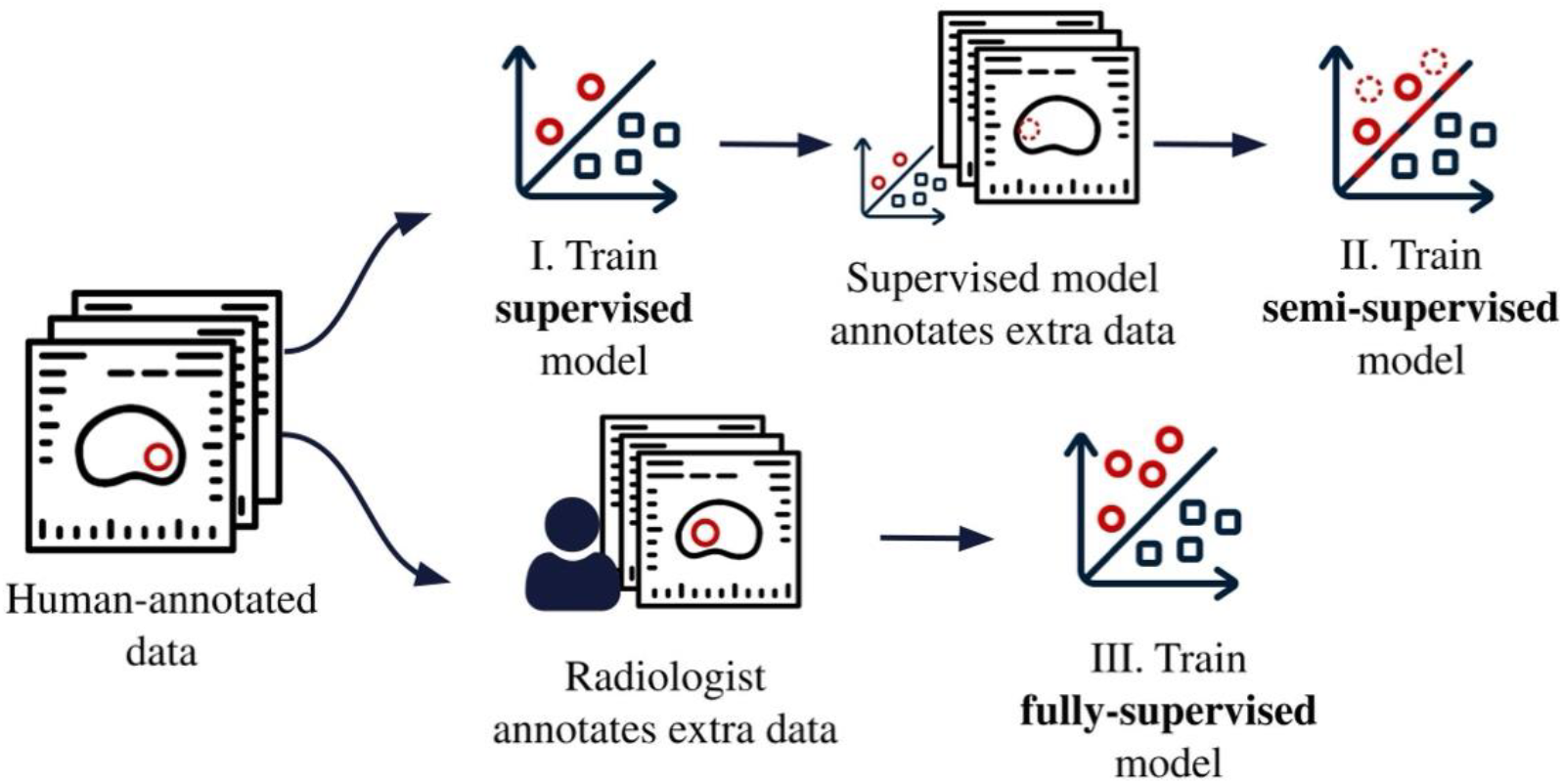
Experimental setup of the three different training strategies.

### 2.4. Model Architecture

The performances of all three strategies were evaluated using three different network architectures. Two well-established frameworks, the nnU-Net [11] and nnDetection [15], and in addition, we propose a new architecture, mtU-Net, as a third semi-supervised approach.

The nnU-Net [11] is a state-of-the-art deep learning framework specifically designed for medical image segmentation tasks. It is based on the U-Net [16] approach for image segmentation and automatically configures data augmentation and architecture based on the specific dataset, often achieving top performance across different benchmarks. The nnDetection [15] is another framework built for medical image analysis inspired by object detection architectures. It is built upon the Retina U-Net [17] architecture, which is a fusion of the Retina Net one-stage object detection approach [18] with the U-Net segmentation architecture [16]. It specializes in detecting objects by using anchor boxes, which are predefined bounding boxes of different sizes and aspect ratios. Concurrently, the network predicts the probability of object inclusion for each anchor box and performs pixel-level segmentation.

### 2.5. mtU-Net

To extend the use of the non-annotated data, we propose mtU-Net, a 3D semi-supervised segmentation model that employs a teacher-student framework integrating pseudo-labeling and consistency regularization. We call our method mtU-Net, based on the Mean Teacher framework [19]. The training strategy consists of using two models with identical architecture, which are denoted student *m*_*s*_ and teacher *m*_*t*_. At every training iteration, both models are fed the same input *x* with a different transformation, and then a consistency loss is computed based on the distance between both models’ predictions (Figure 2).

**Figure 2:**
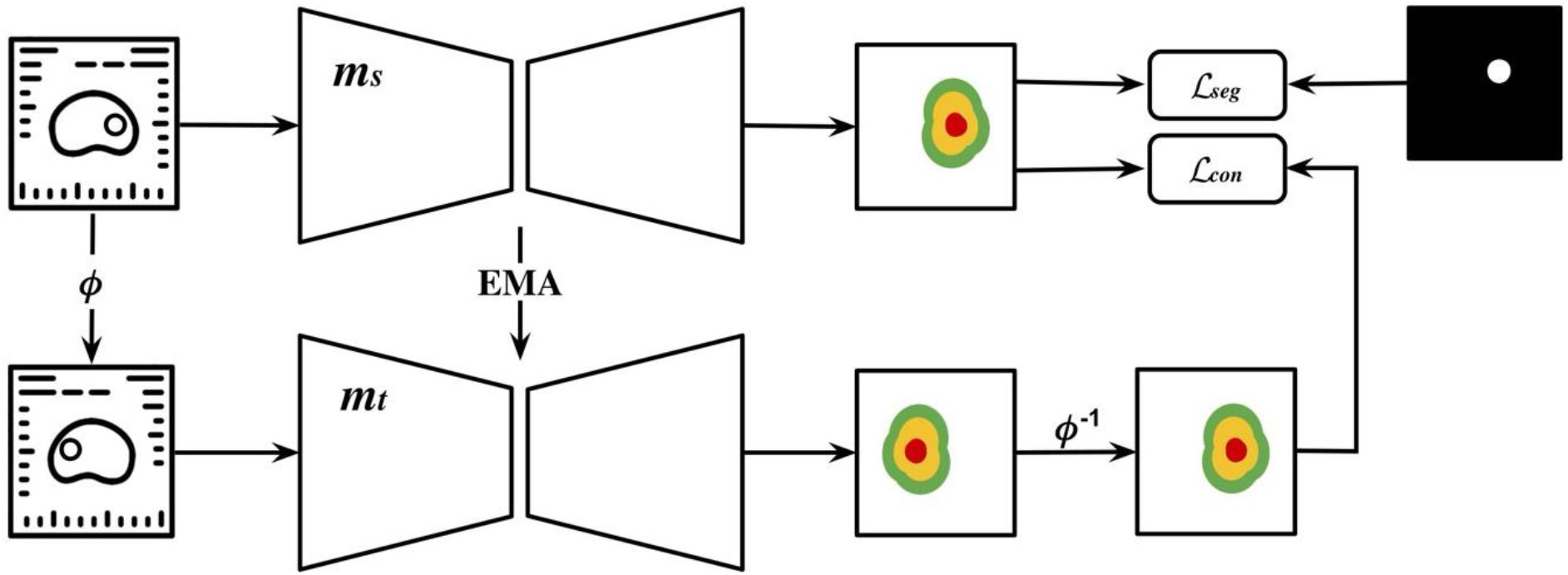
Architecture of mtU-Net. Each sample in the dataset is fed to a student model *m*_*s*_ that outputs segmentation scores, and the segmentation loss ℒ_*seg*_ is computed based on the predictions and ground-truth/pseudo-annotations. The same image receives a transformation *ϕ* and is fed to a teacher model *m*_*t*_, which also generates patch scores. The output of the teacher model receives an inverse transformation *ϕ*^−1^, so the scores match the original image, and the difference between both outputs is added to the training loss as the ℒ_*con*_ to ensure consistency. The student’s weights are updated with backpropagation, and the teacher’s weights are an exponential moving average (EMA) of the student’s weights.

The student weights Θ^*s*^ are updated via loss optimization, and the teacher weights Θ^*t*^ are updated via an exponential moving average (EMA) of the student weights after each training step *e*. A hyperparameter *ρ* controls the EMA decay rate to update the teacher’s weights, as in 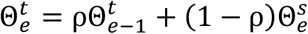.

Consistent with the PI-CAI baseline approach [6], the segmentation loss ℒ_*seg*_ is a combination of focal loss ℒ_*fl*_ [18] with cross-entropy loss ℒ_*ce*_ between the student model’s predictions *m*_*s*_(*x*) and the ground-truth (either human or AI annotations). The focal loss ℒ_*fl*_ = −*α*(1 − *m*_*s*_(*x*))^*γ*^ log(*m*_*s*_(*x*)) ensures a larger weight for the positive class and the less confident predictions by using weighting factor *α* = 0.75 and the focusing parameter *γ* = 2, which is advantageous for segmenting smaller structures such as tumors. Here, *α* balances the importance of positive and negative examples, while *γ* focuses the loss on less-confident examples, thereby improving the model’s ability to accurately segment small lesions. The weight of the focal loss is controlled by a hyperparameter which we choose as *λ*_*fl*_ = 0.5.

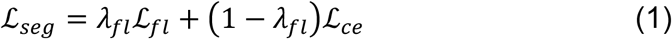

To enforce consistency regularization, we introduce a new loss term to the optimization function. In a classification scenario proposed by [19], the labels remain the same if the input is rotated or shifted; however, in a segmentation task, these functions affect the output, which is a problem when computing the consistency metric. Based on consistency regularization for object detection [20], we propose a straightforward and reversible augmentation strategy denoted as *ϕ*(·), which flips the input image *x* horizontally and, consequently, causes the segmentation output of the model to be flipped. As shown in Equation 2, to compute the consistency loss ℒ_*con*_, we also apply the transformation *ϕ*^−1^(·) to flip the output of the teacher model back to the original input to enforce consistency correctly. A combined loss function ℒ_*comb*_ is used to update the student’s weights. This loss adds the segmentation loss ℒ_*seg*_ to the consistency loss ℒ_*con*_ controlled by a consistency weight hyperparameter *λ*_*con*_.

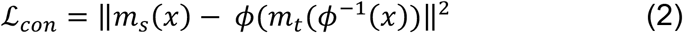

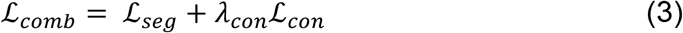

We use the combined loss shown in Equation 3 to update the weights of the mtU-Net student model. The teacher’s weights are updated through an EMA of the student’s weights, controlled by a decay rate hyperparameter we choose as *ρ* = 0.99. All models are trained for a total of 1000 epochs, with a batch size of 3 and patch size of 320×320×16. The consistency weight is controlled by a sigmoid ramp-up function [21] with a length of 300 epochs and a maximum value *λ*_*con*_ = 0.1.

## 3. Results

### 3.1. Annotation comparison

A comparison was performed between the human-annotated and AI-generated lesions within the 205 PI-CAI training samples for which both annotation types were available. Heatmaps illustrate distinct spatial distributions for AI-generated lesions (Figure 3a) compared to human-delineated lesions (Figure 3b). Quantitative analysis revealed differences in spatial coverage and overlap between the annotation types. Human annotations encompassed 83.5% of the total lesion area, whereas AI annotations covered 39.1%. The mean IoU between AI and human annotations was 22.6%. Applying a hit criterion of IoU > 0.10 to assess overlap, the hit ratio between the two annotation sets was 59.9%. Furthermore, the spatial distribution of pathology-aware annotations in the external PROMIS dataset is shown in Figure 3c.

**Figure 3:**
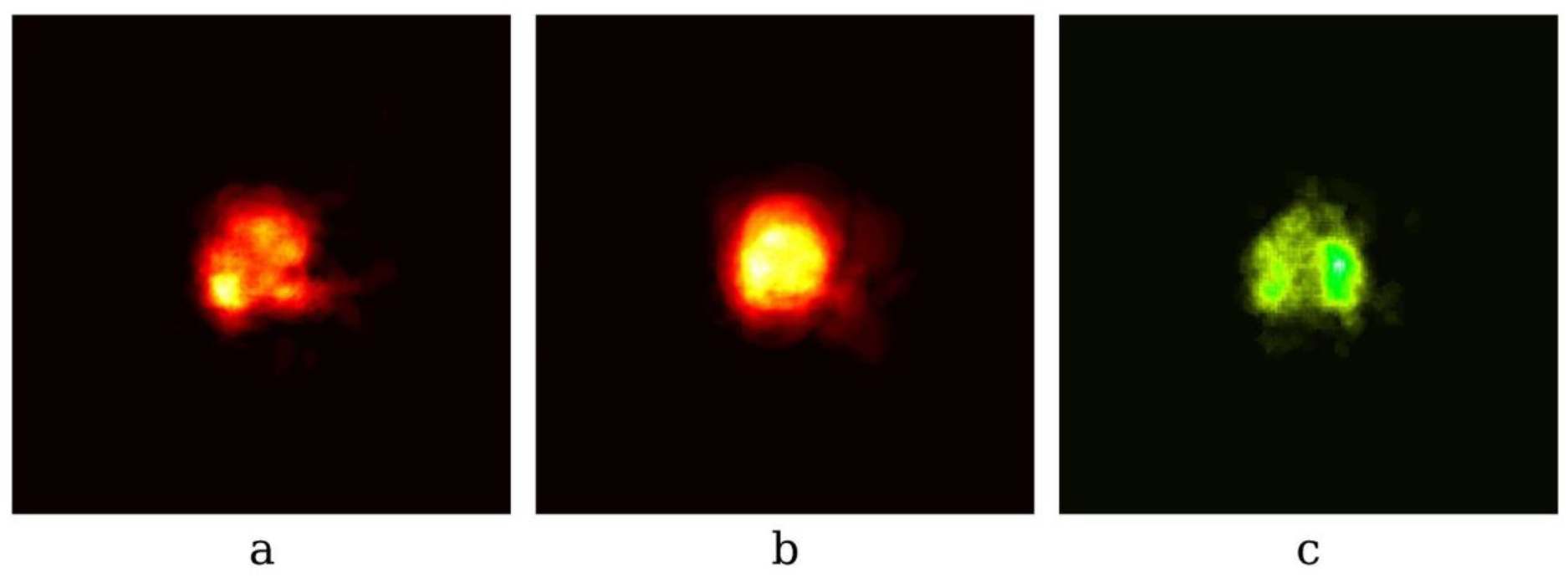
Location heatmap of (a) AI-annotated lesions, (b) human-annotated lesions, and (c) histology-aware annotations from the external validation (PROMIS). Computed on all slices by flattening the Z axis.

### 3.2. Internal model performance

Three deep learning model architectures were investigated: nnU-Net, nnDetection, and mtU-Net. For the nnU-Net and nnDetection architectures, models were trained and compared utilizing supervised, semi-supervised, and fully-supervised strategies. The mtU-Net architecture was evaluated using a semi-supervised training strategy.

**Table 1:** Performance metrics of each training strategy across three test sets. The best results are in bold, and the second best are underlined.

| Architecture | Strategy | PI-CAI Validation<br>(n=100) |  |  | PI-CAI Test<br>(n=1000) |  |  |
| --- | --- | --- | --- | --- | --- | --- | --- |
|  |  | AUC | AP | Avg | AUC | AP | Avg |
| nnU-Net | Supervised | 0.74<br>[0.63-0.83] | 0.46<br>[0.32-0.62] | 0.60<br>[0.48-0.72] | 0.80<br>[0.77-0.83] | 0.45<br>[0.39-0.51] | 0.63<br>[0.59-0.67] |
|  | Semi-supervised | 0.82<br>[0.73-0.90] | 0.61<br>[0.46-0.77] | 0.71<br>[0.61-0.83] | 0.87<br>[0.84-0.89] | <u>0.58</u><br>[0.52-0.63] | <u>0.72</u><br>[0.69-0.76] |
|  | Fully-supervised | **0.89**<br>[0.82-0.94] | **0.73**<br>[0.60-0.84] | **0.81**<br>[0.72-0.89] | **0.89**<br>[0.87-0.91] | **0.65**<br>[0.60-0.70] | **0.77**<br>[0.73-0.80] |
| nnDetection | Supervised | 0.74<br>[0.63-0.83] | 0.27<br>[0.16-0.43] | 0.50<br>[0.41-0.62] | 0.79<br>[0.75-0.82] | 0.39<br>[0.33-0.44] | 0.59<br>[0.55-0.63] |
|  | Semi-supervised | <u>0.89</u><br>[0.81-0.95] | 0.58<br>[0.43-0.74] | 0.73<br>[0.64-0.83] | <u>0.87</u><br>[0.85-0.90] | 0.55<br>[0.49-0.60] | 0.71<br>[0.68-0.75] |
|  | Fully-supervised | 0.80<br>[0.70-0.88] | 0.57<br>[0.43-0.71] | 0.69<br>[0.57-0.79] | 0.83<br>[0.80-0.85] | 0.48<br>[0.43-0.54] | 0.65<br>[0.62-0.69] |
| mtU-Net | Semi-supervised | 0.85<br>[0.77-0.92] | <u>0.68</u><br>[0.55-0.81] | <u>0.77</u><br>[0.67-0.86] | 0.86<br>[0.84-0.89] | 0.58<br>[0.52-0.63] | 0.72<br>[0.68-0.75] |

On the PI-CAI validation set (n=100), the nnU-Net architecture trained with a fully-supervised strategy achieved the highest performance (AUC: 0.89 [95% CI 0.82, 0.94], AP: 0.73 [95% CI 0.60, 0.84]). The semi-supervised strategy showed the second highest performance for the nnU-Net architecture (AUC: 0.82 [95% CI 0.73, 0.90], AP: 0.61 [95% CI 0.46, 0.77]), followed by the nnU-Net baseline supervised strategy (AUC: 0.74 [95% CI 0.63, 0.83], AP: 0.46 [95% CI 0.32, 0.62]). For the nnDetection architecture, the semi-supervised strategy showed the highest results (AUC: 0.89 [95% CI 0.81, 0.95], AP: 0.58 [95% CI 0.43, 0.74]), followed by the fully-supervised (AUC: 0.80 [95% CI 0.70, 0.88], AP: 0.57 [95% CI 0.43, 0.74]), and supervised strategy (AUC: 0.74 [95% CI 0.63, 0.83], AP: 0.27 [95% CI 0.16, 0.43]), which presented the lowest AP over all models in the dataset. The mtU-Net semi-supervised strategy showed higher AUC and AP (AUC: 0.85 [95% CI 0.77, 0.92], AP: 0.68 [95% CI 0.55, 0.81]) than the semi-supervised nnU-Net (AUC: 0.82 [95% CI 0.73, 0.90], AP: 0.61 [95% CI 0.46, 0.77]), on this dataset.

On the PI-CAI test set (n=1000), the nnU-Net architecture trained with a fully-supervised strategy also achieved the highest performance (AUC: 0.89 [95% CI 0.87, 0.91], AP: 0.65 [95% CI 0.60,0.70]). The semi-supervised strategy was the second highest performance of the nnU-Net architecture (AUC: 0.87 [95% CI 0.84, 0.89], AP: 0.58 [95% CI 0.52, 0.63]) followed by the supervised nnU-Net (AUC: 0.80 [95% CI 0.77, 0.83], AP: 0.45 [95% CI 0.39, 0.51]). For the nnDetection architecture, the semi-supervised strategy showed the highest results (AUC: 0.87 [95% CI 0.85, 0.90], AP: 0.55 [95% CI 0.49, 0.60]), followed by the fully-supervised (AUC: 0.83 [95% CI 0.80, 0.85], AP: 0.48 [95% CI 0.43, 0.54]), and supervised (AUC: 0.79 [95% CI 0.75, 0.82], AP: 0.39 [95% CI 0.33, 0.44]), which was the lowest overall performance. In contrast to the validation set, in the PI-CAI test set the mtU-Net semi-supervised strategy (AUC: 0.86 [95% CI 0.84, 0.89], AP: 0.58 [95% CI 0.52, 0.63]) shows numerically close performance metrics to the nnU-net semi-supervised strategy (AUC: 0.87 [95% CI 0.84, 0.89], AP: 0.58 [95% CI 0.52, 0.63]).

### 3.3. External model performance

On the PROMIS external validation set (n=574) the comparative performance of the different strategies is visualized by the ROC curves in Figure 4a. Within the nnU-Net architecture group, the fully-supervised strategy achieved the highest performance with a significantly higher AUC than the supervised baseline (AUC: 0.70 [95% CI 0.66, 0.74], AP: 0.24 [95% CI 0.19, 0.29]; p = 0.002). The semi-supervised nnU-Net strategy also demonstrated a significantly higher AUC compared to the supervised baseline (AUC: 0.65 [95% CI 0.60, 0.69], AP: 0.20 [95% CI 0.16, 0.24]; p = 0.042). The nnU-Net baseline supervised strategy yielded an AUC of 0.61 [95% CI 0.57, 0.66] and AP of 0.16 [95% CI 0.12, 0.20]. For the nnDetection architecture, neither the fully-supervised strategy (AUC: 0.60 [95% CI 0.55, 0.64], AP: 0.10 [95% CI 0.06, 0.13]; p = 0.256) nor the semi-supervised strategy (AUC: 0.59 [95% CI 0.55, 0.64], AP: 0.13 [95% CI 0.09, 0.16]; p = 0.123) showed a statistically significant difference in AUC compared to the nnDetection supervised baseline (AUC: 0.56 [95% CI 0.52, 0.61], AP: 0.12 [95% CI 0.09, 0.16]). The mtU-Net semi-supervised strategy achieved the second highest overall metrics, with an AUC significantly higher than the nnU-Net supervised baseline (AUC: 0.66 [95% CI 0.62, 0.71], AP: 0.20 [95% CI 0.16, 0.25]; p = 0.024).

**Figure 4:**
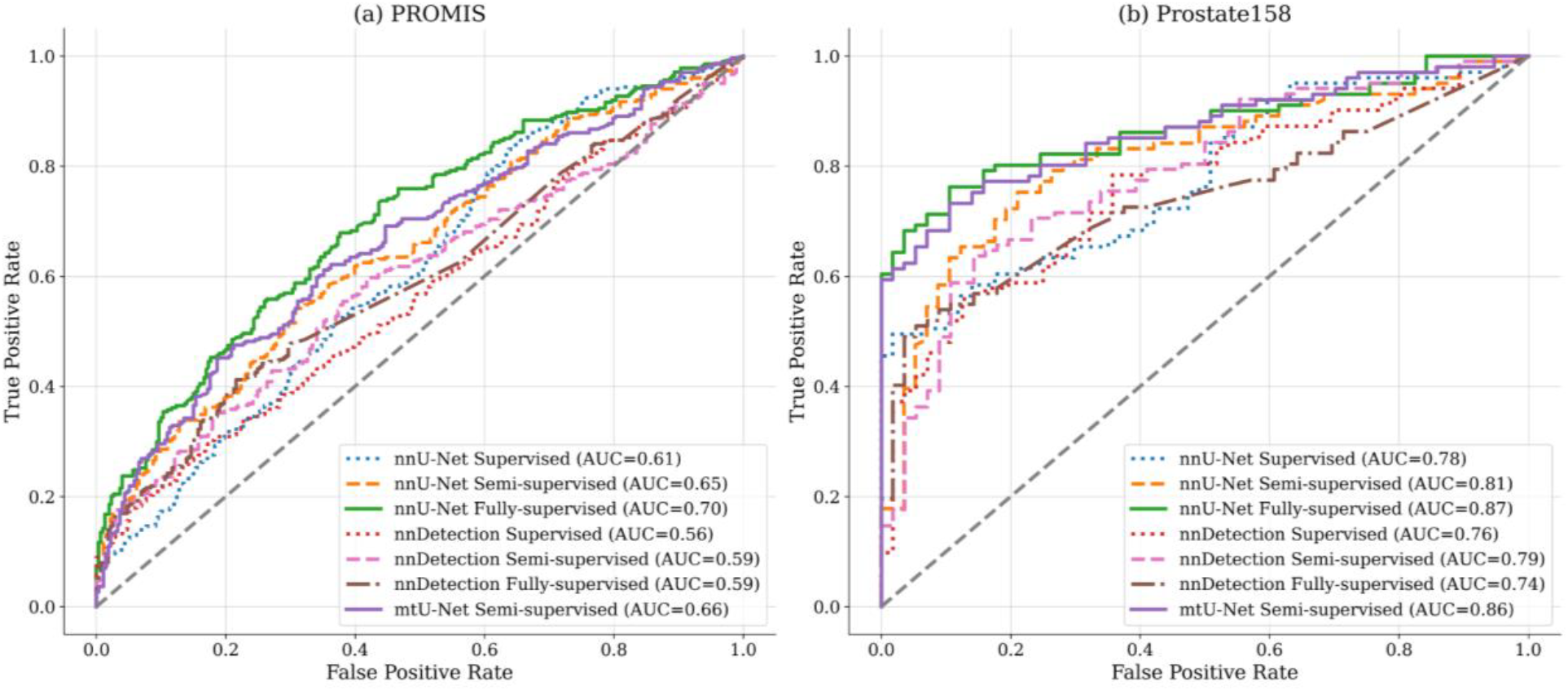
ROC curve comparison of models across the external validation datasets (a) PROMIS and (b) Prostate158.

On the Prostate158 external validation set (n=158), Figure 4b displays the ROC curves for each approach. The nnU-Net fully-supervised strategy again achieved the highest performance, with a significantly higher AUC compared to the supervised baseline (AUC: 0.87 [95% CI 0.82, 0.92], AP: 0.64 [95% CI 0.56, 0.72]; p = 0.002). The nnU-Net semi-supervised strategy did not show a statistically significant AUC difference from the supervised baseline (AUC: 0.81 [95% CI 0.74, 0.88], AP: 0.53 [95% CI 0.44, 0.62]; p = 0.256). The nnU-Net baseline supervised strategy achieved an AUC of 0.78 [95% CI 0.71, 0.84] and AP of 0.41 [95% CI 0.32, 0.51]. Within the nnDetection architecture group, neither the semi-supervised strategy (AUC: 0.79 [95% CI 0.72, 0.86], AP: 0.44 [95% CI 0.35, 0.53]; p = 0.360) nor the fully-supervised strategy (AUC: 0.74 [95% CI 0.67, 0.81], AP: 0.38 [95% CI 0.29, 0.47]; p = 0.561) demonstrated a significant AUC difference compared to the nnDetection supervised baseline (AUC: 0.76 [95% CI 0.68, 0.83], AP: 0.45 [95% CI 0.36, 0.54]). The mtU-Net semi-supervised strategy showed the second highest metrics over all architectures, with an AUC significantly higher than the nnU-Net supervised baseline (AUC: 0.86 [95% CI 0.81, 0.92], AP: 0.58 [95% CI 0.49, 0.67]; p = 0.007), and numerically close to the top-performing fully-supervised nnU-Net.

**Table 2:** AUC, AP, and 95% Confidence Intervals for different models across PROMIS and P158 datasets. P-values from the DeLong test comparing model performance against supervised baselines. The best results are in bold, and the second best are underlined.

| Architecture | Strategy | PROMIS<br>($n=574$) | | | Prostate158<br>($n=158$) | | |
| --- | --- | --- | --- | --- | --- | --- | --- |
| | | AUC | AP | $p <$ | AUC | AP | $p <$ |
| nnU-Net | Supervised | 0.61<br>[0.57-0.66] | 0.16<br>[0.12-0.20] | - | 0.78<br>[0.71-0.84] | 0.41<br>[0.32-0.51] | - |
|  | Semi-supervised | 0.65<br>[0.60-0.69] | 0.20<br>[0.16-0.24] | **0.042** | 0.81<br>[0.74-0.88] | 0.53<br>[0.44-0.62] | 0.256 |
|  | Fully-supervised | **0.70**<br>[0.66-0.74] | **0.24**<br>[0.19-0.29] | **0.002** | **0.87**<br>[0.82-0.92] | **0.64**<br>[0.56-0.72] | **0.002** |
| nnDetection | Supervised | 0.56<br>[0.52-0.61] | 0.12<br>[0.09-0.16] | - | 0.76<br>[0.68-0.83] | 0.45<br>[0.36-0.54] | - |
|  | Semi-supervised | 0.59<br>[0.55-0.64] | 0.13<br>[0.09-0.16] | 0.123 | 0.79<br>[0.72-0.86] | 0.44<br>[0.35-0.53] | 0.360 |
|  | Fully-supervised | 0.60<br>[0.55-0.64] | 0.10<br>[0.06-0.13] | 0.256 | 0.74<br>[0.67-0.81] | 0.38<br>[0.29-0.47] | 0.561 |
| mtU-Net | Semi-supervised | <u>0.66</u><br>[0.62-0.71] | <u>0.20</u><br>[0.16-0.25] | **0.024** | <u>0.86</u><br>[0.81-0.92] | <u>0.58</u><br>[0.49-0.67] | **0.007** |

## 4. Discussion

This study aimed to provide a comprehensive evaluation of semi-supervised learning for prostate cancer segmentation and to determine its viability as an alternative to the addition of more human-annotated data. The findings contribute to understanding the potential of SSL approaches in clinical applications where high-quality annotations are limited.

While a fully-supervised model showed the highest performance internally, semi-supervised approaches, particularly our proposed mtU-Net, offered comparable performance on external datasets, approaching the fully-supervised benchmark. Our analysis of training strategies revealed that fully-supervised learning with nnU-Net yielded the highest performance on internal validation datasets, demonstrating a clear advantage of maximizing human-annotated, high quality data. However, on external validation datasets, the performance of semi-supervised approaches closely approached that of the fully-supervised model, aligning with the findings of previous studies [12, 21]. This observation suggests that while extensive human annotation may provide a performance boost in internal evaluations, its benefits might not fully generalize to external datasets. The marginal improvements seen in fully-supervised models on external data, compared to semi-supervised methods, suggest that increasing efforts in human annotation of internal data might not proportionally enhance the external performance of the model. This implies that prioritizing efforts to increase data diversity using semi-supervised training approaches could potentially yield more robust and generalizable models.

By integrating consistency regularization with pseudo-labeling, our proposed mtU-Net architecture was specifically designed to mitigate the impact of noise from AI-generated annotations, leading to improved model generalizability. This was evidenced by its consistently strong performance across the internal PI-CAI test sets and external PROMIS and Prostate158 datasets, and the only semi-supervised approach that showed statistically significant improvement over the supervised baseline on Prostate158 and closely aligned ROC curves with the fully-supervised model. The effectiveness of this approach underscores that architectural suitability varies, not all architectures handle different learning strategies or potentially inaccurate AI-generated annotations equally well. For example, nnDetection demonstrated this variability, with metrics indicating comparative underperformance in fully-supervised settings relative to semi-supervised applications, suggesting that a lesser dependency on high-quality annotations might benefit certain model types.

We observed differences between the human and AI annotations, specifically the larger surface area and broader distribution of human-annotated lesions versus the localized and conservative nature of AI annotations. This suggests that AI models, when trained mostly on AI-annotated data, may prioritize precision over recall, potentially reducing sensitivity. The conservative annotation style on the AI-annotated data, focusing on smaller areas with higher probability of aggressive cancer, indicates a need for careful consideration when using such data for training, particularly in clinical applications where sensitivity is crucial. Furthermore, the distinct lesion distribution observed in the PROMIS samples underscores that AI models are subject not only to technical variability in bpMRI acquisition but also to variability between populations, and ground-truth annotation. This highlights the importance of validating models on diverse external datasets to ensure generalizability.

This study has potential limitations. The 205 manual annotations used for the fully-supervised learning were delineated by one radiologist without access to histopathology reports and inter-reader variability was not assessed. The AI-generated annotations were based on a specific methodology [12] and the effectiveness of semi-supervised learning might differ depending on the quality and characteristics of pseudo-annotations generated by other techniques.

In conclusion, adding expert annotations improved internal performance but offered limited gains in generalizability compared to semi-supervised learning. Semi-supervised models achieved similar external validation performance to fully-supervised learning by using non-annotated data, highlighting that data volume and diversity, not just expert annotations, drive robustness. This supports semi-supervised learning as a valuable and viable approach for aggressive prostate tumor segmentation, maintaining competitive, generalizable performance while reducing the need for extensive costly expert annotations.

## Data Availability

All data produced in the present study are available upon reasonable request to the authors

## References

[1] F. Bray, J. Ferlay, I. Soerjomataram, R. L. Siegel, L. A. Torre, A. Jemal, Global cancer statistics 2018: GLOBOCAN estimates of incidence and mortality worldwide for 36 cancers in 185 countries, CA Cancer J. Clin. 68 (2018) 394–424.

[2] F. Giganti, A. B. Rosenkrantz, G. Villeirs, V. Panebianco, A. Stabile, M. Emberton, C. M. Moore, The evolution of mri of the prostate: the past, the present, and the future, American Journal of Roentgenology 213 (2019) 384–396.

[3] E. Bass, A. Pantovic, M. Connor, R. Gabe, A. Padhani, A. Rockall, H. Sokhi, H. Tam, M. Winkler, H. Ahmed, A systematic review and meta-analysis of the diagnostic accuracy of biparametric prostate mri for prostate cancer in men at risk, Prostate Cancer and Prostatic Diseases 24 (2021) 596–611.

[4] A. C. Westphalen, C. E. McCulloch, J. M. Anaokar, S. Arora, N. S. Barashi, J. O. Barentsz, T. K. Bathala, L. K. Bittencourt, M. T. Booker, V. G. Braxton, P. R. Carroll, D. D. Casalino, S. D. Chang, F. V. Coakley, R. Dhatt, S. C. Eberhardt, B. R. Foster, A. T. Froemming, J. J. Fütterer, D. M. Ganeshan, M. R. Gertner, L. Mankowski Gettle, S. Ghai, R. T. Gupta, M. E. Hahn, R. Houshyar, C. Kim, C. K. Kim, C. Lall, D. J. A. Margolis, S. E. McRae, A. Oto, R. B. Parsons, N. U. Patel, P. A. Pinto, T. J. Polascik, B. Spilseth, J. B. Starcevich, V. S. Tammisetti, S. S. Taneja, B. Turkbey, S. Verma, J. F. Ward, C. A. Warlick, A. R. Weinberger, J. Yu, R. J. Zagoria, A. B. Rosenkrantz, Variability of the positive predictive value of PI-RADS for prostate MRI across 26 centers: Experience of the society of abdominal radiology prostate cancer disease-focused panel, Radiology 296 (2020) 76–84.

[5] S. J. C. Soerensen, R. E. Fan, A. Seetharaman, L. Chen, W. Shao, I. Bhattacharya, Y.-H. Kim, R. Sood, M. Borre, B. I. Chung, K. J. To’o, M. Rusu, G. A. Sonn, Deep learning improves speed and accuracy of prostate gland segmentations on magnetic resonance imaging for targeted biopsy, J. Urol. 206 (2021) 604–612.

[6] A. Saha, J. Bosma, J. Twilt, B. van Ginneken, D. Yakar, M. Elschot, J. Veltman, J. Fütterer, M. de Rooij, H. Huisman, Artificial intelligence and radiologists at prostate cancer detection in MRI — the PI-CAI challenge, in: Medical Imaging with Deep Learning, short paper track, 2023.

[7] D.-H. Lee, Pseudo-Label: The simple and efficient Semi-Supervised learning method for deep neural networks, Workshop on challenges in representation learning, ICML 3 (2013).

[8] X. Luo, M. Hu, T. Song, G. Wang, S. Zhang, Semi-supervised medical image segmentation via cross teaching between cnn and transformer, in: International Conference on Medical Imaging with Deep Learning, PMLR, 2022, pp. 820–833.

[9] X. Luo, J. Chen, T. Song, G. Wang, Semi-supervised medical image segmentation through dual-task consistency, 2021, pp. 8801–8809.

[10] X. Luo, G. Wang, W. Liao, J. Chen, T. Song, Y. Chen, D. N. M. Zhang, Shichuan, S. Zhang, Semi-supervised medical image segmentation via uncertainty rectified pyramid consistency, Medical Image Analysis 80 (2022) 102517.

[11] F. Isensee, P. F. Jaeger, S. A. A. Kohl, J. Petersen, K. H. Maier-Hein, nnU-Net: a self-configuring method for deep learning-based biomedical image segmentation, Nat. Methods 18 (2020) 203–211.

[12] J. S. Bosma, A. Saha, M. Hosseinzadeh, I. Slootweg, M. de Rooij, H. Huisman, Semisupervised learning with report-guided pseudo labels for deep learning–based prostate cancer detection using biparametric MRI, Radiology: Artificial Intelligence (2023).

[13] H. U. Ahmed, A. E.-S. Bosaily, L. C. Brown, R. Gabe, R. Kaplan, M. K. Parmar, Y. Collaco-Moraes, K. Ward, R. G. Hindley, A. Freeman, et al., Diagnostic accuracy of multi-parametric mri and trus biopsy in prostate cancer (promis): a paired validating confirmatory study, The Lancet 389 (2017) 815–822.

[14] L. C. Adams, M. R. Makowski, G. Engel, M. Rattunde, F. Busch, P. Asbach, S. M. Niehues, S. Vinayahalingam, B. van Ginneken, G. Litjens, K. K. Bressem, Prostate158 - an expert-annotated 3t mri dataset and algorithm for prostate cancer detection, Computers in Biology and Medicine 148 (2022) 105817. M. Baumgartner, P. F. Jäger, F. Isensee, K. H. Maier-Hein, nndetection: A self-configuring method for medical object detection, Medical Image Computing and Computer Assisted Intervention – MICCAI 2021 (2021) 530–539.

[15] O. Ronneberger, P. Fischer, T. Brox, U-Net: Convolutional networks for biomedical image segmentation, Med. Image Comput. Comput. Assist. Interv. (2015) 234–241.

[16] P. Jaeger, S. A. A. Kohl, S. Bickelhaupt, F. Isensee, T. Kuder, H. Schlemmer, K. Maier-Hein, Retina U-Net: Embarrassingly simple exploitation of segmentation supervision for medical object detection, ML4H@NeurIPS (2018).

[17] T.-Y. Lin, P. Goyal, R. Girshick, K. He, P. Dollár, Focal loss for dense object detection, in: Proceedings of the IEEE international conference on computer vision, 2017, pp. 2980–2988.

[18] A. Tarvainen, H. Valpola, Mean teachers are better role models: Weight-averaged consistency targets improve semi-supervised deep learning results, in: Annual Conference on Neural Information Processing Systems, 2017, pp. 1195–1204.

[19] J. Jeong, S. Lee, J. Kim, N. Kwak, Consistency-based semi-supervised learning for object detection, in: Annual Conference on Neural Information Processing Systems, 2019, pp. 10758–10767.

[20] S. Laine, T. Aila, Temporal ensembling for semisupervised learning, CoRR abs/1610.02242 (2016). URL:http://arxiv.org/abs/1610.02242. arXiv:1610.02242.

[21] Singh P, Chukkapalli R, Chaudhari S, Chen L, Chen M, Pan J, Smuda C, Cirrone J. Shifting to machine supervision: annotation-efficient semi and self-supervised learning for automatic medical image segmentation and classification. Sci Rep. 2024 May 11;14(1):10820. doi: 10.1038/s41598-024-61822-9. PMID: 38734825; PMCID: PMC11088676.

